# Nonlinear associations between obesity indices and fall risk in middle-aged and older Chinese adults: a cross-sectional analysis based on CHARLS

**DOI:** 10.1101/2025.08.01.25332621

**Authors:** Zihao Liu, Jiahe Zhang, Yongjun Li

## Abstract

**Background:** Falls are a leading cause of disability and mortality among older adults in China. The mechanisms linking various obesity indices—such as BMI, Waist-to-Height Ratio (WHtR), Body Roundness Index (BRI), and Body Fat Percentage (BFP) — to fall risk remain unclear. Given the limitations of BMI, it is important to investigate the influence of body composition and central obesity indicators on fall risk.

**Methods:** This cross-sectional study used data from the China Health and Retirement Longitudinal Study (CHARLS, 2011–2016; n = 27,303). Logistic regression, restricted cubic spline (RCS) modeling, and piecewise regression were applied to explore the nonlinear associations between obesity indices and falls. Covariates including demographic and health-related factors were adjusted for in multivariate models.

**Results:** WHtR, BRI, and BFP were nonlinearly associated with fall risk, while BMI showed no significant relationship. Inflection points were observed at 0.504 for WHtR, 3.06 for BRI, and 28.6% for BFP. Below these thresholds, increases in WHtR and BFP were associated with reduced fall risk; above the thresholds, fall risk increased significantly. Subgroup analyses indicated stronger associations among women, rural residents, and individuals with arthritis.

**Conclusion:** Our findings identify WHtR, BRI, and BFP as more sensitive predictors of fall risk than BMI. Thresholds of BRI ≥ 3.0 and BFP ≥ 28% may serve as clinical screening markers for fall prevention, supporting a shift from BMI-centric to integrated body composition-function assessments.

## Introduction

Falls pose a serious public health challenge among the elderly population. They result in significant injury, morbidity, and mortality rates, and place enormous economic pressure on health care systems [1]. Statistics show that falls have become one of the leading causes of accidental injury and death among people aged 60 and older, with both the frequency of falls and the severity of related complications on the rise [2]. In China, for example, about 50 million elderly people experience at least one fall per year. Among them, 36% to 44% require emergency medical attention due to falls. These individuals are also at higher risk of falling again or even dying within a year [3]. Studies have shown an important association between obesity and fall risk, especially in older adults, where obesity not only affects balance and coordination, but also leads to reduced exercise capacity. Moreover, obesity may increase the burden on the lower limbs, thereby affecting gait and stability in older adults [4, 5]. A study in Nigeria found that 77.5% of obese patients reported at least one fall in the past year, significantly higher than 17.2% of non-obese patients [6]. In addition, studies have shown that obesity and physical inactivity together increase the risk of falls, although this effect is not always statistically significant [7]. In a Canadian study, obese men had a significantly higher risk of falls than men of other weight categories, but this increased risk was not evident in women [8]. Therefore, in-depth exploration of the relationship between obesity indicators and fall risk is crucial for formulating more effective fall prevention strategies, improving the quality of life of the elderly, and reducing the negative effects of falls such as disability and social isolation [9]. To sum up, exploring the risk factors and protective factors of falls and implementing appropriate interventions have become necessary strategies to address this increasingly serious public health problem.

Obesity measurement indices are important tools to assess overweight and obesity in individuals. These indices include Body Mass Index (BMI), Waist-to-Height Ratio (WHtR), Body Roundness Index (BRI), and Body Fat Percentage (BFP). They assess body fat content and distribution by measuring weight, height, and specific body circumferences. BMI is a widely used measure of obesity, although it has significant limitations: it does not distinguish individuals with similar BMI values but different body fat percentages [10].However, when analyzing correlations between disease risk factors and obesity measures, differences in body composition and fat distribution must be considered. Specifically, BFP reflects body composition, while WHtR indicates abdominal fat accumulation and changes in fat distribution. This information helps healthcare professionals and individuals assess health risks and develop strategies for disease prevention and health improvement. Several studies have explored the relationship between these indicators and falls. For example, one study found that dynamic stability and balance decreased as BMI increased in older adults [12]. Another observational study of 15,860 older adults in an Italian community reported that underweight and obese individuals fell more often than those with normal or overweight status, with a stronger effect observed in men than women [13]. Furthermore, another study noted that both high BMI and low grip strength were associated with a higher risk of falls [14]. These findings underscore the importance of weight management in fall prevention. However, the underlying mechanisms of the interaction between obesity and falls remain inadequately explained. Moreover, there is a lack of studies examining the relationship between obesity and falls in the Chinese population. Therefore, this study aims to explore the association between various obesity measures and fall events among middle-aged and elderly people in China.

The China Longitudinal Study of Health and Retirement (CHARLS) is a nationally representative, multi-dimensional, longitudinal project that maintains continuity through the organization’s annual survey of Chinese population samples [15]. A recent analysis based on CHARLS data reveals significant regional differences in the incidence of falls among middle-aged and elderly adults in China. Although the northwest region has the lowest risk of falls, it ranks first in the incidence of hip fractures nationwide, suggesting that the distribution of body composition may play a modulating role [16]. It should be noted that existing studies have not systematically examined the nonlinear association between obesity parameters and falls in the Chinese population, nor accounted for potential confounding factors. This study highlights the need to investigate risk factors affecting the incidence of falls in older adults in China. Therefore, CHARLS provides a nationally representative, high-quality sample to examine the relationship between obesity measures and falls.

In this study, we aim to explore the relationship between obesity measures and falls in a national sample of the Chinese population. Ultimately, we seek to provide scientific evidence for fall prevention strategies in the elderly. Clarifying these relationships provides guidance for clinical practice and public health interventions aimed at reducing fall-related events in this vulnerable population.

## Materials and methods

### Data and study populations

CHARLS data were collected and screened for cross-sectional studies. CHARLS is a biennial, nationally representative longitudinal survey conducted by the China Center for Economic Research at Peking University. With assistance from 28 provincial-level Centers for Disease Control and Prevention (CDC), the CHARLS Beijing Office first gathered lists and contact information of county-level CDC personnel. Before census takers arrived at the three selected villages or communities to conduct mapping, county-level CDC liaison officers communicated with local leaders to facilitate coordination between census takers, village or community heads, and residents.

Ethical approval for the original CHARLS project was granted by the Peking University Biomedical Ethics Committee (IRB00001052–11015), and all participants provided written informed consent. Our current work is a secondary analysis of the fully anonymized, publicly available CHARLS data. We combined data from CHARLS waves conducted in 2011-2012, 2013-2014, and 2015-2016.

All participants underwent structured face-to-face family interviews using comprehensive questionnaires. CHARLS employs a multistage probability sampling strategy incorporating the Probability Proportional to Size (PPS) technique. This method includes four stages: county sampling, community-level sampling, household-level sampling, and interviewee-level sampling to ensure a nationally representative sample.

During district sampling, 150 districts representing 28 provinces were selected. At the community level, rural villages and urban communities served as primary sampling units (PSUs), with three PSUs randomly chosen from each county. Households were then selected based on detailed maps and lists from public service units. Finally, an individual aged 45 or older was randomly selected from each household as the primary respondent [15].

### Fall events and obesity index measures

The outcome variable of this study was whether participants had a fall event in the past two years, identified based on participants’ self-reports in the CHARLS questionnaires: "Have you fallen in the past two years?" If the participant answered "yes," they were classified as having fallen [17]. The same doctor measured all participants, including height, weight, and waist circumference, and calculated obesity measures. Body mass index (BMI) was calculated as weight (kg) divided by height (m) squared; waist-to-height ratio (WHtR) was calculated as waist circumference divided by height; body roundness index (BRI) was defined as follows: BRI = 364.2 – 365.5 × √[1 – (WC/(2π))2/(0.5 × height)2]. Body fat percentage (BFP) was calculated using the following formulas: for adult males, BFP = 1.20 × BMI + 0.23 × age − 16.2; for adult females, BFP = 1.20 × BMI + 0.23 × age − 5.4 [18,19]. Height was measured using a Seca (TM) 213 rangefinder (Medical Scale and Measurement System Saika (Hangzhou) Co., Ltd.), weight was measured using an Omron (TM) HN-286 scale (Krel Precision (Yangzhou) Co., Ltd.), and waist circumference was measured using a flexible tape measure.

### Covariate identification for cross-sectional studies

The covariates in this cross-sectional study were divided into three categories. The first category, demographic characteristics, included sex, age (continuous variable), marital status (married, single, divorced, widowed), residence (rural or urban), and education level (primary school and below, junior high school, senior high school, and above). The second category, lifestyle factors, comprised smoking status (never, former, or current), alcohol consumption (never, former, or current), vision correction (use of glasses: yes or no), and hearing impairment, which was self-rated on a five-category scale (no impairment, mild, moderate, severe, profound). The third category consisted of chronic diseases, including hypertension, diabetes, dyslipidemia, heart disease, stroke, chronic lung disease, asthma, chronic kidney disease, gastric disease, arthritis, memory disorders, mental disorders, cancer, and disability. All covariates were obtained from structured questionnaires.

### Statistic analysis

Continuous variables were expressed as means with 95% confidence intervals (CIs), and categorical variables as counts (percentages). The relationships between obesity measures and falls were analyzed using logistic regression. We also applied restricted cubic spline (RCS) regression to examine the nonlinear association between obesity measures and falls. Subgroup analyses by sex were performed to assess whether significant interactions existed between sex and the associations of obesity measures with falls. Odds ratios (ORs) with 95% CIs and P-values were calculated. All analyses were performed using the Free Statistics software version 2.1. A P-value less than 0.05 was considered statistically significant, using a two-tailed test.

## Results

### Cross-sectional study participant characteristics

According to the China Longitudinal Study of Health and Retirement (2011–2016), 55,144 participants were initially enrolled. After excluding those with no record of falls (N=1,740), incomplete obesity measurements (N=12,388), and extreme obesity indices (defined as values below the 0.5th percentile or above the 99.5th percentile, N=13,713), a total of 27,303 participants were included in the final analysis (Figure 1). Of these, 4,696 participants (17.2%) reported falls. As shown in Table 1, participants who experienced falls were older (62.0 ± 9.9 vs. 60.2 ± 9.7 years, P < 0.001), had a higher proportion of females (60% vs. 51.2%, P < 0.001), and were more likely to live in rural areas (64.1% vs. 62.4%, P = 0.024).Significant differences were observed between the fall group and the non-fall group in the prevalence of chronic diseases such as hypertension (36% vs. 30.3%, P < 0.001), diabetes (11.1% vs. 8.1%, P < 0.001), and arthritis (51.1% vs. 37.1%, P < 0.001). Obesity measures,including waist-to-height ratio (WHtR), body roundness index (BRI), and body fat percentage (BFP), were higher in the fall group (WHtR: 0.6 ± 0.1 vs. 0.5 ± 0.1, P < 0.001; BRI: 4.5 ± 1.5 vs. 4.4 ± 1.4, P < 0.001; BFP: 33.1 ± 7.4 vs. 31.8 ± 7.2, P < 0.001), while body mass index (BMI) did not differ significantly between the two groups. The population characteristics remained generally consistent before and after multiple imputation for missing data. This consistency further supports the robustness of the observed associations (Table 1, column after imputation).

**Fig 1.**
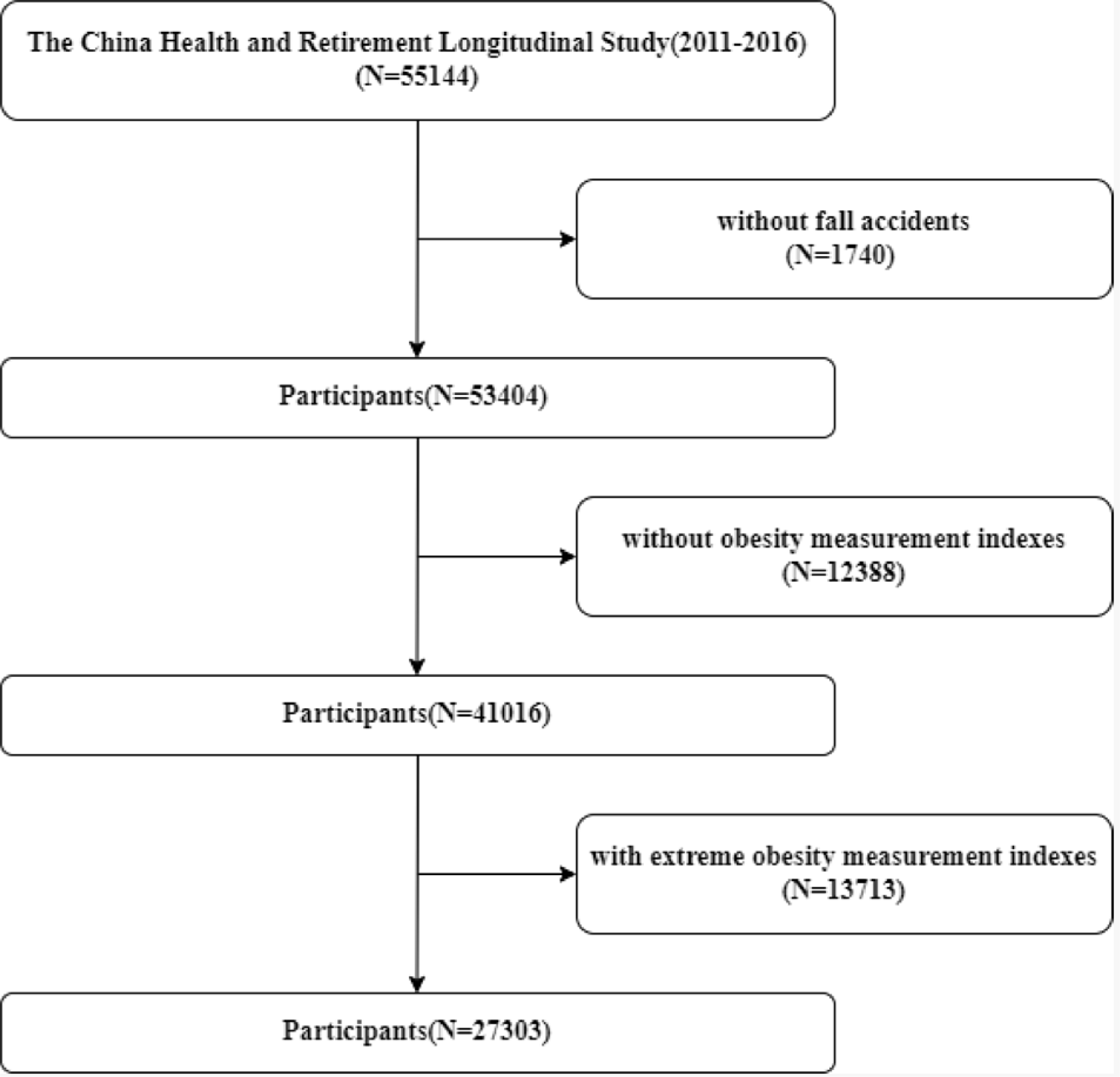
Flowchart depicting the participants’ selection

**Table 1.** General characteristics of participants. Crude model adjusted for:none. Model 1 adjusted for:sex, age, marital status, residence, and education level;. Model 2 adjusted for: sex, age, marital status, residence, education level, drink, smoke, vision correction and hearing impairment.Model 3 adjusted for: sex, age, marital status, residence, education level, drink, smoke, vision correction, hearing impairment, hypertension, diabetes, dyslipidemia, heart disease, stroke, chronic lung disease, asthma, chronic kidney disease, gastric disease, arthritis, memory disorders, mental disorders, cancer and disability.

| Variables | Before Multiple Imputation |  |  |  |  | After Multiple Imputation |  |  |  |  |
| --- | --- | --- | --- | --- | --- | --- | --- | --- | --- | --- |
|  | Total<br>(n=27303) | No Fall<br>(n=22607) | Fall<br>(n=4696) | Statistic | P | Total<br>(n= 27303) | No Fall<br>(n=22607) | Fall<br>(n=4696) | Statistic | P |
| **Sex** |  |  |  | 121.025 | < 0.001 |  |  |  | 121.025 | < 0.001 |
| Female | 14387 (52.7) | 11570 (51.2) | 2817 (60) |  |  | 14387 (52.7) | 11570 (51.2) | 2817 (60) |  |  |
| Male | 12916 (47.3) | 11037 (48.8) | 1879 (40) |  |  | 12916 (47.3) | 11037 (48.8) | 1879 (40) |  |  |
| **Marital status** |  |  |  | 58.628 | < 0.001 |  |  |  | 58.7 | < 0.001 |
| Others(unmarried<br>/divorced/widowed) | 3580 (13.1) | 2803 (12.4) | 777 (16.5) |  |  | 3580 (13.1) | 2803 (12.4) | 777 (16.5) |  |  |
| Married | 23719 (86.9) | 19800 (87.6) | 3919 (83.5) |  |  | 23723 (86.9) | 19804 (87.6) | 3919 (83.5) |  |  |
| **Residence place** |  |  |  | 5.077 | 0.024 |  |  |  | 5.077 | 0.024 |
| Urban | 10186 (37.3) | 8502 (37.6) | 1684 (35.9) |  |  | 10186 (37.3) | 8502 (37.6) | 1684 (35.9) |  |  |
| Rural | 17117 (62.7) | 14105 (62.4) | 3012 (64.1) |  |  | 17117 (62.7) | 14105 (62.4) | 3012 (64.1) |  |  |
| **Age** | 60.5 ± 9.7 | 60.2 ± 9.7 | 62.0 ± 9.9 | 130.806 | < 0.001 | 60.5 ± 9.7 | 60.2 ± 9.7 | 62.0 ± 9.9 | 130.806 | < 0.001 |
| **Education** |  |  |  | 119.986 | < 0.001 |  |  |  | 120.298 | < 0.001 |
| Primary school or below | 12132 (44.5) | 9736 (43.1) | 2396 (51) |  |  | 12139 (44.5) | 9743 (43.1) | 2396 (51) |  |  |
| Primary school | 6874 (25.2) | 5730 (25.4) | 1144 (24.4) |  |  | 6879 (25.2) | 5735 (25.4) | 1144 (24.4) |  |  |
| Secondary school | 5363 (19.7) | 4588 (20.3) | 775 (16.5) |  |  | 5369 (19.7) | 4594 (20.3) | 775 (16.5) |  |  |
| High school or above | 2913 (10.7) | 2532 (11.2) | 381 (8.1) |  |  | 2916 (10.7) | 2535 (11.2) | 381 (8.1) |  |  |
| **Drink** |  |  |  | 0.699 | 0.403 |  |  |  | 0.731 | 0.393 |
| Never drinker | 14818 (54.4) | 12297 (54.5) | 2521 (53.8) |  |  | 14835 (54.3) | 12310 (54.5) | 2525 (53.8) |  |  |
| Ever drinker (past or<br>current) | 12445 (45.6) | 10280 (45.5) | 2165 (46.2) |  |  | 12468 (45.7) | 10297 (45.5) | 2171 (46.2) |  |  |
| **Smoke** |  |  |  | 22.433 | < 0.001 |  |  |  | 26.609 | < 0.001 |
| Never smoker | 15599 (63.0) | 12733 (62.3) | 2866 (66.1) |  |  | 16310 (59.7) | 13347 (59) | 2963 (63.1) |  |  |
| Ever smoker (past or<br>current) | 9178 (37.0) | 7709 (37.7) | 1469 (33.9) |  |  | 10993 (40.3) | 9260 (41) | 1733 (36.9) |  |  |
| **Hypertension** |  |  |  | 54.962 | < 0.001 |  |  |  | 52.282 | < 0.001 |
| No | 17056 (68.7) | 14292 (69.7) | 2764 (64) |  |  | 18891 (69.2) | 15850 (70.1) | 3041 (64.8) |  |  |
| Yes | 7764 (31.3) | 6207 (30.3) | 1557 (36) |  |  | 8412 (30.8) | 6757 (29.9) | 1655 (35.2) |  |  |
| **Diabetes** |  |  |  | 38.667 | < 0.001 |  |  |  | 44.906 | < 0.001 |
| No | 22550 (91.4) | 18738 (91.9) | 3812 (88.9) |  |  | 24919 (91.3) | 20751 (91.8) | 4168 (88.8) |  |  |
| Yes | 2131 ( 8.6) | 1657 (8.1) | 474 (11.1) |  |  | 2384 ( 8.7) | 1856 (8.2) | 528 (11.2) |  |  |
| **Cancer** |  |  |  | 0.416 | 0.519 |  |  |  | 0.256 | 0.613 |
| No | 24473 (98.6) | 20212 (98.6) | 4261 (98.7) |  |  | 26921 (98.6) | 22287 (98.6) | 4634 (98.7) |  |  |
| Yes | 348 ( 1.4) | 292 (1.4) | 56 (1.3) |  |  | 382 ( 1.4) | 320 (1.4) | 62 (1.3) |  |  |
| **Pulmonary Disease** |  |  |  | 57.872 | < 0.001 |  |  |  | 71.085 | < 0.001 |
| No | 21671 (87.2) | 18057 (88) | 3614 (83.7) |  |  | 23843 (87.3) | 19917 (88.1) | 3926 (83.6) |  |  |
| Yes | 3173 (12.8) | 2470 (12) | 703 (16.3) |  |  | 3460 (12.7) | 2690 (11.9) | 770 (16.4) |  |  |
| **Heart Disease** |  |  |  | 77.089 | < 0.001 |  |  |  | 77.364 | < 0.001 |
| No | 20830 (84.0) | 17407 (84.9) | 3423 (79.5) |  |  | 22963 (84.1) | 19214 (85) | 3749 (79.8) |  |  |
| Yes | 3970 (16.0) | 3089 (15.1) | 881 (20.5) |  |  | 4340 (15.9) | 3393 (15) | 947 (20.2) |  |  |
| **Stroke** |  |  |  | 75.463 | < 0.001 |  |  |  | 88.713 | < 0.001 |
| No | 24096 (96.7) | 20004 (97.2) | 4092 (94.6) |  |  | 26419 (96.8) | 21979 (97.2) | 4440 (94.5) |  |  |
| Yes | 811 ( 3.3) | 578 (2.8) | 233 (5.4) |  |  | 884 ( 3.2) | 628 (2.8) | 256 (5.5) |  |  |
| **Mental Disorders** |  |  |  | 34.001 | < 0.001 |  |  |  | 45.598 | < 0.001 |
| No | 24409 (98.2) | 20208 (98.4) | 4201 (97.1) |  |  | 26805 (98.2) | 22251 (98.4) | 4554 (97) |  |  |
| Yes | 451 ( 1.8) | 326 (1.6) | 125 (2.9) |  |  | 498 ( 1.8) | 356 (1.6) | 142 (3) |  |  |
| **Arthritis** |  |  |  | 294.058 | < 0.001 |  |  |  | 304.678 | < 0.001 |
| No | 15058 (60.5) | 12938 (62.9) | 2120 (48.9) |  |  | 16546 (60.6) | 14232 (63) | 2314 (49.3) |  |  |
| Yes | 9846 (39.5) | 7630 (37.1) | 2216 (51.1) |  |  | 10757 (39.4) | 8375 (37) | 2382 (50.7) |  |  |
| **Dyslipidemia** |  |  |  | 26.636 | < 0.001 |  |  |  | 30.466 | < 0.001 |
| No | 20303 (83.9) | 16916 (84.4) | 3387 (81.2) |  |  | 22883 (83.8) | 19074 (84.4) | 3809 (81.1) |  |  |
| Yes | 3908 (16.1) | 3123 (15.6) | 785 (18.8) |  |  | 4420 (16.2) | 3533 (15.6) | 887 (18.9) |  |  |
| **Liver Disease** |  |  |  | 37.554 | < 0.001 |  |  |  | 46.51 | < 0.001 |
| No | 23388 (94.5) | 19416 (94.9) | 3972 (92.5) |  |  | 25753 (94.3) | 21422 (94.8) | 4331 (92.2) |  |  |
| Yes | 1365 ( 5.5) | 1045 (5.1) | 320 (7.5) |  |  | 1550 ( 5.7) | 1185 (5.2) | 365 (7.8) |  |  |
| **Kidney Disease** |  |  |  | 95.557 | < 0.001 |  |  |  | 100.787 | < 0.001 |
| No | 22688 (91.5) | 18912 (92.3) | 3776 (87.8) |  |  | 24981 (91.5) | 20859 (92.3) | 4122 (87.8) |  |  |
| Yes | 2096 ( 8.5) | 1570 (7.7) | 526 (12.2) |  |  | 2322 ( 8.5) | 1748 (7.7) | 574 (12.2) |  |  |
| **Gastrointestinal Disease** |  |  |  | 169.512 | < 0.001 |  |  |  | 175.047 | < 0.001 |
| No | 17833 (71.6) | 15086 (73.3) | 2747 (63.5) |  |  | 19499 (71.4) | 16518 (73.1) | 2981 (63.5) |  |  |
| Yes | 7064 (28.4) | 5486 (26.7) | 1578 (36.5) |  |  | 7804 (28.6) | 6089 (26.9) | 1715 (36.5) |  |  |
| **Asthma** |  |  |  | 26.18 | < 0.001 |  |  |  | 33.051 | < 0.001 |
| No | 23533 (94.6) | 19514 (94.9) | 4019 (93) |  |  | 25862 (94.7) | 21494 (95.1) | 4368 (93) |  |  |
| Yes | 1341 ( 5.4) | 1039 (5.1) | 302 (7) |  |  | 1441 ( 5.3) | 1113 (4.9) | 328 (7) |  |  |
| **Memory Disorders** |  |  |  | 46.81 | < 0.001 |  |  |  | 50.17 | < 0.001 |
| No | 24286 (97.7) | 20121 (98) | 4165 (96.3) |  |  | 26676 (97.7) | 22154 (98) | 4522 (96.3) |  |  |
| Yes | 577 ( 2.3) | 415 (2) | 162 (3.7) |  |  | 627 ( 2.3) | 453 (2) | 174 (3.7) |  |  |
| **Disability** |  |  |  | 348.89 | < 0.001 |  |  |  | 364.904 | < 0.001 |
| No | 19256 (74.8) | 16461 (77.1) | 2795 (63.7) |  |  | 20232 (74.1) | 17274 (76.4) | 2958 (63) |  |  |
| Yes | 6483 (25.2) | 4888 (22.9) | 1595 (36.3) |  |  | 7071 (25.9) | 5333 (23.6) | 1738 (37) |  |  |
| **Glasses Use** |  |  |  | 5.179 | 0.075 |  |  |  | 5.177 | 0.075 |
| No | 19648 (72.0) | 16331 (72.3) | 3317 (70.6) |  |  | 19653 (72.0) | 16336 (72.3) | 3317 (70.6) |  |  |
| Yes | 7588 (27.8) | 6219 (27.5) | 1369 (29.2) |  |  | 7590 (27.8) | 6221 (27.5) | 1369 (29.2) |  |  |
| Blind | 60 ( 0.2) | 50 (0.2) | 10 (0.2) |  |  | 60 ( 0.2) | 50 (0.2) | 10 (0.2) |  |  |
| **Hearing Impairment** |  |  |  | 248.873 | < 0.001 |  |  |  | 257.905 | < 0.001 |
| Poor | 3558 (13.4) | 2701 (12.2) | 857 (18.8) |  |  | 3754 (13.7) | 2847 (12.6) | 907 (19.3) |  |  |
| Fair | 13250 (49.8) | 10841 (49.2) | 2409 (52.9) |  |  | 13577 (49.7) | 11101 (49.1) | 2476 (52.7) |  |  |
| Good | 5051 (19.0) | 4358 (19.8) | 693 (15.2) |  |  | 5151 (18.9) | 4441 (19.6) | 710 (15.1) |  |  |
| Very good | 4135 (15.5) | 3605 (16.3) | 530 (11.6) |  |  | 4200 (15.4) | 3659 (16.2) | 541 (11.5) |  |  |
| Excellent | 607 ( 2.3) | 546 (2.5) | 61 (1.3) |  |  | 621 ( 2.3) | 559 (2.5) | 62 (1.3) |  |  |
| **BMI(kg/m^2)** | 23.9 ± 3.6 | 23.9 ± 3.5 | 23.8 ± 3.6 | 0.886 | 0.347 | 23.9 ± 3.6 | 23.9 ± 3.5 | 23.8 ± 3.6 | 0.886 | 0.347 |
| **WHtR<sup>a</sup>** | 0.6 ± 0.1 | 0.5 ± 0.1 | 0.6 ± 0.1 | 43.538 | < 0.001 | 0.6 ± 0.1 | 0.5 ± 0.1 | 0.6 ± 0.1 | 43.538 | < 0.001 |
| **BRI<sup>b</sup>** | 4.4 ± 1.4 | 4.4 ± 1.4 | 4.5 ± 1.5 | 26.335 | < 0.001 | 4.4 ± 1.4 | 4.4 ± 1.4 | 4.5 ± 1.5 | 26.335 | < 0.001 |
| **BFP<sup>c</sup>** | 32.0 ± 7.2 | 31.8 ± 7.2 | 33.1 ± 7.4 | 126.086 | < 0.001 | 32.0 ± 7.2 | 31.8 ± 7.2 | 33.1 ± 7.4 | 126.086 | < 0.001 |

<sup>a</sup>Waist to height ratio.
<sup>b</sup>Body fat percentage.
<sup>c</sup>Body roundness index.

**Table 1 General characteristics of participants**
| Variables | Before Multiple Imputation |  |  |  |  | After Multiple Imputation |  |  |  |  |
| --- | --- | --- | --- | --- | --- | --- | --- | --- | --- | --- |
|  | Total<br>(n=27303) | No Fall<br>(n=22607) | Fall<br>(n=4696) | Statistic | P | Total<br>(n= 27303) | No Fall<br>(n=22607) | Fall<br>(n=4696) | Statistic | P |
| **Sex** |  |  |  | 121.025 | < 0.001 |  |  |  | 121.025 | < 0.001 |
| Female | 14387 (52.7) | 11570 (51.2) | 2817 (60) |  |  | 14387 (52.7) | 11570 (51.2) | 2817 (60) |  |  |
| Male | 12916 (47.3) | 11037 (48.8) | 1879 (40) |  |  | 12916 (47.3) | 11037 (48.8) | 1879 (40) |  |  |
| **Marital status** |  |  |  | 58.628 | < 0.001 |  |  |  | 58.7 | < 0.001 |
| Others(unmarried<br>/divorced/widowed) | 3580 (13.1) | 2803 (12.4) | 777 (16.5) |  |  | 3580 (13.1) | 2803 (12.4) | 777 (16.5) |  |  |
| Married | 23719 (86.9) | 19800 (87.6) | 3919 (83.5) |  |  | 23723 (86.9) | 19804 (87.6) | 3919 (83.5) |  |  |
| **Residence place** |  |  |  | 5.077 | 0.024 |  |  |  | 5.077 | 0.024 |
| Urban | 10186 (37.3) | 8502 (37.6) | 1684 (35.9) |  |  | 10186 (37.3) | 8502 (37.6) | 1684 (35.9) |  |  |
| Rural | 17117 (62.7) | 14105 (62.4) | 3012 (64.1) |  |  | 17117 (62.7) | 14105 (62.4) | 3012 (64.1) |  |  |
| **Age** | 60.5 ± 9.7 | 60.2 ± 9.7 | 62.0 ± 9.9 | 130.806 | < 0.001 | 60.5 ± 9.7 | 60.2 ± 9.7 | 62.0 ± 9.9 | 130.806 | < 0.001 |
| **Education** |  |  |  | 119.986 | < 0.001 |  |  |  | 120.298 | < 0.001 |
| Primary school or below | 12132 (44.5) | 9736 (43.1) | 2396 (51) |  |  | 12139 (44.5) | 9743 (43.1) | 2396 (51) |  |  |
| Primary school | 6874 (25.2) | 5730 (25.4) | 1144 (24.4) |  |  | 6879 (25.2) | 5735 (25.4) | 1144 (24.4) |  |  |
| Secondary school | 5363 (19.7) | 4588 (20.3) | 775 (16.5) |  |  | 5369 (19.7) | 4594 (20.3) | 775 (16.5) |  |  |
| High school or above | 2913 (10.7) | 2532 (11.2) | 381 (8.1) |  |  | 2916 (10.7) | 2535 (11.2) | 381 (8.1) |  |  |
| **Drink** |  |  |  | 0.699 | 0.403 |  |  |  | 0.731 | 0.393 |
| Never drinker | 14818 (54.4) | 12297 (54.5) | 2521 (53.8) |  |  | 14835 (54.3) | 12310 (54.5) | 2525 (53.8) |  |  |
| Ever drinker (past or<br>current) | 12445 (45.6) | 10280 (45.5) | 2165 (46.2) |  |  | 12468 (45.7) | 10297 (45.5) | 2171 (46.2) |  |  |
| **Smoke** |  |  |  | 22.433 | < 0.001 |  |  |  | 26.609 | < 0.001 |
| Never smoker | 15599 (63.0) | 12733 (62.3) | 2866 (66.1) |  |  | 16310 (59.7) | 13347 (59) | 2963 (63.1) |  |  |
| Ever smoker (past or current) | 9178 (37.0) | 7709 (37.7) | 1469 (33.9) |  |  | 10993 (40.3) | 9260 (41) | 1733 (36.9) |  |  |
| **Hypertension** |  |  |  | 54.962 | < 0.001 |  |  |  | 52.282 | < 0.001 |
| No | 17056 (68.7) | 14292 (69.7) | 2764 (64) |  |  | 18891 (69.2) | 15850 (70.1) | 3041 (64.8) |  |  |
| Yes | 7764 (31.3) | 6207 (30.3) | 1557 (36) |  |  | 8412 (30.8) | 6757 (29.9) | 1655 (35.2) |  |  |
| **Diabetes** |  |  |  | 38.667 | < 0.001 |  |  |  | 44.906 | < 0.001 |
| No | 22550 (91.4) | 18738 (91.9) | 3812 (88.9) |  |  | 24919 (91.3) | 20751 (91.8) | 4168 (88.8) |  |  |
| Yes | 2131 ( 8.6) | 1657 (8.1) | 474 (11.1) |  |  | 2384 ( 8.7) | 1856 (8.2) | 528 (11.2) |  |  |
| **Cancer** |  |  |  | 0.416 | 0.519 |  |  |  | 0.256 | 0.613 |
| No | 24473 (98.6) | 20212 (98.6) | 4261 (98.7) |  |  | 26921 (98.6) | 22287 (98.6) | 4634 (98.7) |  |  |
| Yes | 348 ( 1.4) | 292 (1.4) | 56 (1.3) |  |  | 382 ( 1.4) | 320 (1.4) | 62 (1.3) |  |  |
| **Pulmonary Disease** |  |  |  | 57.872 | < 0.001 |  |  |  | 71.085 | < 0.001 |
| No | 21671 (87.2) | 18057 (88) | 3614 (83.7) |  |  | 23843 (87.3) | 19917 (88.1) | 3926 (83.6) |  |  |
| Yes | 3173 (12.8) | 2470 (12) | 703 (16.3) |  |  | 3460 (12.7) | 2690 (11.9) | 770 (16.4) |  |  |
| **Heart Disease** |  |  |  | 77.089 | < 0.001 |  |  |  | 77.364 | < 0.001 |
| No | 20830 (84.0) | 17407 (84.9) | 3423 (79.5) |  |  | 22963 (84.1) | 19214 (85) | 3749 (79.8) |  |  |
| Yes | 3970 (16.0) | 3089 (15.1) | 881 (20.5) |  |  | 4340 (15.9) | 3393 (15) | 947 (20.2) |  |  |
| **Stroke** |  |  |  | 75.463 | < 0.001 |  |  |  | 88.713 | < 0.001 |
| No | 24096 (96.7) | 20004 (97.2) | 4092 (94.6) |  |  | 26419 (96.8) | 21979 (97.2) | 4440 (94.5) |  |  |
| Yes | 811 ( 3.3) | 578 (2.8) | 233 (5.4) |  |  | 884 ( 3.2) | 628 (2.8) | 256 (5.5) |  |  |
| **Mental Disorders** |  |  |  | 34.001 | < 0.001 |  |  |  | 45.598 | < 0.001 |
| No | 24409 (98.2) | 20208 (98.4) | 4201 (97.1) |  |  | 26805 (98.2) | 22251 (98.4) | 4554 (97) |  |  |
| Yes | 451 ( 1.8) | 326 (1.6) | 125 (2.9) |  |  | 498 ( 1.8) | 356 (1.6) | 142 (3) |  |  |
| **Arthritis** |  |  |  | 294.058 | < 0.001 |  |  |  | 304.678 | < 0.001 |
| No | 15058 (60.5) | 12938 (62.9) | 2120 (48.9) |  |  | 16546 (60.6) | 14232 (63) | 2314 (49.3) |  |  |
| Yes | 9846 (39.5) | 7630 (37.1) | 2216 (51.1) |  |  | 10757 (39.4) | 8375 (37) | 2382 (50.7) |  |  |
| **Dyslipidemia** |  |  |  | 26.636 | < 0.001 |  |  |  | 30.466 | < 0.001 |
| No | 20303 (83.9) | 16916 (84.4) | 3387 (81.2) |  |  | 22883 (83.8) | 19074 (84.4) | 3809 (81.1) |  |  |
| Yes | 3908 (16.1) | 3123 (15.6) | 785 (18.8) |  |  | 4420 (16.2) | 3533 (15.6) | 887 (18.9) |  |  |
| **Liver Disease** |  |  |  | 37.554 | < 0.001 |  |  |  | 46.51 | < 0.001 |
| No | 23388 (94.5) | 19416 (94.9) | 3972 (92.5) |  |  | 25753 (94.3) | 21422 (94.8) | 4331 (92.2) |  |  |
| Yes | 1365 ( 5.5) | 1045 (5.1) | 320 (7.5) |  |  | 1550 ( 5.7) | 1185 (5.2) | 365 (7.8) |  |  |
| **Kidney Disease** |  |  |  | 95.557 | < 0.001 |  |  |  | 100.787 | < 0.001 |
| No | 22688 (91.5) | 18912 (92.3) | 3776 (87.8) |  |  | 24981 (91.5) | 20859 (92.3) | 4122 (87.8) |  |  |
| Yes | 2096 ( 8.5) | 1570 (7.7) | 526 (12.2) |  |  | 2322 ( 8.5) | 1748 (7.7) | 574 (12.2) |  |  |
| **Gastrointestinal Disease** |  |  |  | 169.512 | < 0.001 |  |  |  | 175.047 | < 0.001 |
| No | 17833 (71.6) | 15086 (73.3) | 2747 (63.5) |  |  | 19499 (71.4) | 16518 (73.1) | 2981 (63.5) |  |  |
| Yes | 7064 (28.4) | 5486 (26.7) | 1578 (36.5) |  |  | 7804 (28.6) | 6089 (26.9) | 1715 (36.5) |  |  |
| **Asthma** |  |  |  | 26.18 | < 0.001 |  |  |  | 33.051 | < 0.001 |
| No | 23533 (94.6) | 19514 (94.9) | 4019 (93) |  |  | 25862 (94.7) | 21494 (95.1) | 4368 (93) |  |  |
| Yes | 1341 ( 5.4) | 1039 (5.1) | 302 (7) |  |  | 1441 ( 5.3) | 1113 (4.9) | 328 (7) |  |  |
| **Memory Disorders** |  |  |  | 46.81 | < 0.001 |  |  |  | 50.17 | < 0.001 |
| No | 24286 (97.7) | 20121 (98) | 4165 (96.3) |  |  | 26676 (97.7) | 22154 (98) | 4522 (96.3) |  |  |
| Yes | 577 ( 2.3) | 415 (2) | 162 (3.7) |  |  | 627 ( 2.3) | 453 (2) | 174 (3.7) |  |  |
| **Disability** |  |  |  | 348.89 | < 0.001 |  |  |  | 364.904 | < 0.001 |
| No | 19256 (74.8) | 16461 (77.1) | 2795 (63.7) |  |  | 20232 (74.1) | 17274 (76.4) | 2958 (63) |  |  |
| Yes | 6483 (25.2) | 4888 (22.9) | 1595 (36.3) |  |  | 7071 (25.9) | 5333 (23.6) | 1738 (37) |  |  |
| **Glasse Use** |  |  |  | 5.179 | 0.075 |  |  |  | 5.177 | 0.075 |
| No | 19648 (72.0) | 16331 (72.3) | 3317 (70.6) |  |  | 19653 (72.0) | 16336 (72.3) | 3317 (70.6) |  |  |
| Yes | 7588 (27.8) | 6219 (27.5) | 1369 (29.2) |  |  | 7590 (27.8) | 6221 (27.5) | 1369 (29.2) |  |  |
| Blind | 60 ( 0.2) | 50 (0.2) | 10 (0.2) |  |  | 60 ( 0.2) | 50 (0.2) | 10 (0.2) |  |  |
| **Hearing Impairment** |  |  |  | 248.873 | < 0.001 |  |  |  | 257.905 | < 0.001 |
| Poor | 3558 (13.4) | 2701 (12.2) | 857 (18.8) |  |  | 3754 (13.7) | 2847 (12.6) | 907 (19.3) |  |  |
| Fair | 13250 (49.8) | 10841 (49.2) | 2409 (52.9) |  |  | 13577 (49.7) | 11101 (49.1) | 2476 (52.7) |  |  |
| Good | 5051 (19.0) | 4358 (19.8) | 693 (15.2) |  |  | 5151 (18.9) | 4441 (19.6) | 710 (15.1) |  |  |
| Very good | 4135 (15.5) | 3605 (16.3) | 530 (11.6) |  |  | 4200 (15.4) | 3659 (16.2) | 541 (11.5) |  |  |
| Excellent | 607 ( 2.3) | 546 (2.5) | 61 (1.3) |  |  | 621 ( 2.3) | 559 (2.5) | 62 (1.3) |  |  |
| **BMI(kg/m<sup>2</sup>)** | 23.9 ± 3.6 | 23.9 ± 3.5 | 23.8 ± 3.6 | 0.886 | 0.347 | 23.9 ± 3.6 | 23.9 ± 3.5 | 23.8 ± 3.6 | 0.886 | 0.347 |
| **WHtR<sup>a</sup>** | 0.6 ± 0.1 | 0.5 ± 0.1 | 0.6 ± 0.1 | 43.538 | < 0.001 | 0.6 ± 0.1 | 0.5 ± 0.1 | 0.6 ± 0.1 | 43.538 | < 0.001 |
| **BRI<sup>b</sup>** | 4.4 ± 1.4 | 4.4 ± 1.4 | 4.5 ± 1.5 | 26.335 | < 0.001 | 4.4 ± 1.4 | 4.4 ± 1.4 | 4.5 ± 1.5 | 26.335 | < 0.001 |
| **BFP<sup>c</sup>** | 32.0 ± 7.2 | 31.8 ± 7.2 | 33.1 ± 7.4 | 126.086 | < 0.001 | 32.0 ± 7.2 | 31.8 ± 7.2 | 33.1 ± 7.4 | 126.086 | < 0.001 |

<sup>a</sup>Waist to height ratio.
<sup>b</sup>Body fat percentage.
<sup>c</sup>Body roundness index.

### Association between obesity markers and falls

A logistic regression model was developed to analyze the relationship between measures of obesity and falls (Table 2). The results revealed differences in the effects of various categorical variable quantiles. In the univariate crude model, BMI as a continuous variable was not significantly associated with falls (OR = 1, 95% CI 0.99–1, P = 0.347). Similarly, its categorical variables, including the highest quantile BMI-Q3 (OR = 1, 95% CI 0.93–1.08, P = 0.937) and the lowest quantile BMI-Q1 (OR = 1.05, 95% CI 0.97–1.13, P = 0.217), were not statistically significant. WHtR was significantly associated as a continuous variable (OR = 1.02, P < 0.001), with the highest quartile WHtR-Q3 showing a stronger association (OR = 1.22, 95% CI 1.13–1.32, P < 0.001). BRI as a continuous variable (OR = 1.01, P < 0.001) and the highest quartile BRI-Q3 (OR = 1.21, 95% CI 1.12–1.31, P < 0.001) were significant in univariate models, while BFP was also significant as a continuous variable (OR = 1.03, P < 0.001) and the highest quartile BFP-Q3 (OR = 1.41, 95% CI 1.31–1.52, P < 0.001). After adjusting for sex, age, and other variables in model 1, WHtR-Q3 remained significant (OR = 1.11, 95% CI 1.02–1.20, P = 0.011). The effect size for BRI-Q3 decreased slightly but remained significant (OR = 1.10, 95% CI 1.02–1.19, P = 0.015). Similarly, the OR for BFP-Q3 decreased but stayed significant (OR = 1.16, 95% CI 1.06–1.27, P = 0.001). Compared with model 1, further adjustment in model 2 strengthened the associations of WHtR-Q3 (OR = 1.12, 95% CI 1.04–1.22, P = 0.003), BRI-Q3 (OR = 1.12, 95% CI 1.04–1.21, P = 0.005), and BFP-Q3 (OR = 1.18, 95% CI 1.08–1.29, P < 0.001). In the final model 3, WHtR lost significance as a continuous variable (OR = 1, P = 0.068), which may be due to the adjustment of confounding factors affecting the linear association; however, the highest quartiles of WHtR-Q3 (OR = 1.09, 95% CI 1.01–1.18, P = 0.036), BRI-Q3 (OR = 1.08, 95% CI 1.00–1.17, P = 0.049), and BFP-Q3 (OR = 1.15, 95% CI 1.05–1.26, P = 0.002) remained statistically significant and robust. The lowest quartiles (e.g., BMI-Q1, WHtR-Q1) showed no significant association in any model. The results indicated that the highest quartiles of obesity (e.g., WHtR-Q3, BRI-Q3, BFP-Q3) were independent risk factors for falls, with odds ratios remaining significant after multiple adjustments. In contrast, the middle (reference) and lowest quartiles showed no significant association.

**Table 2.**
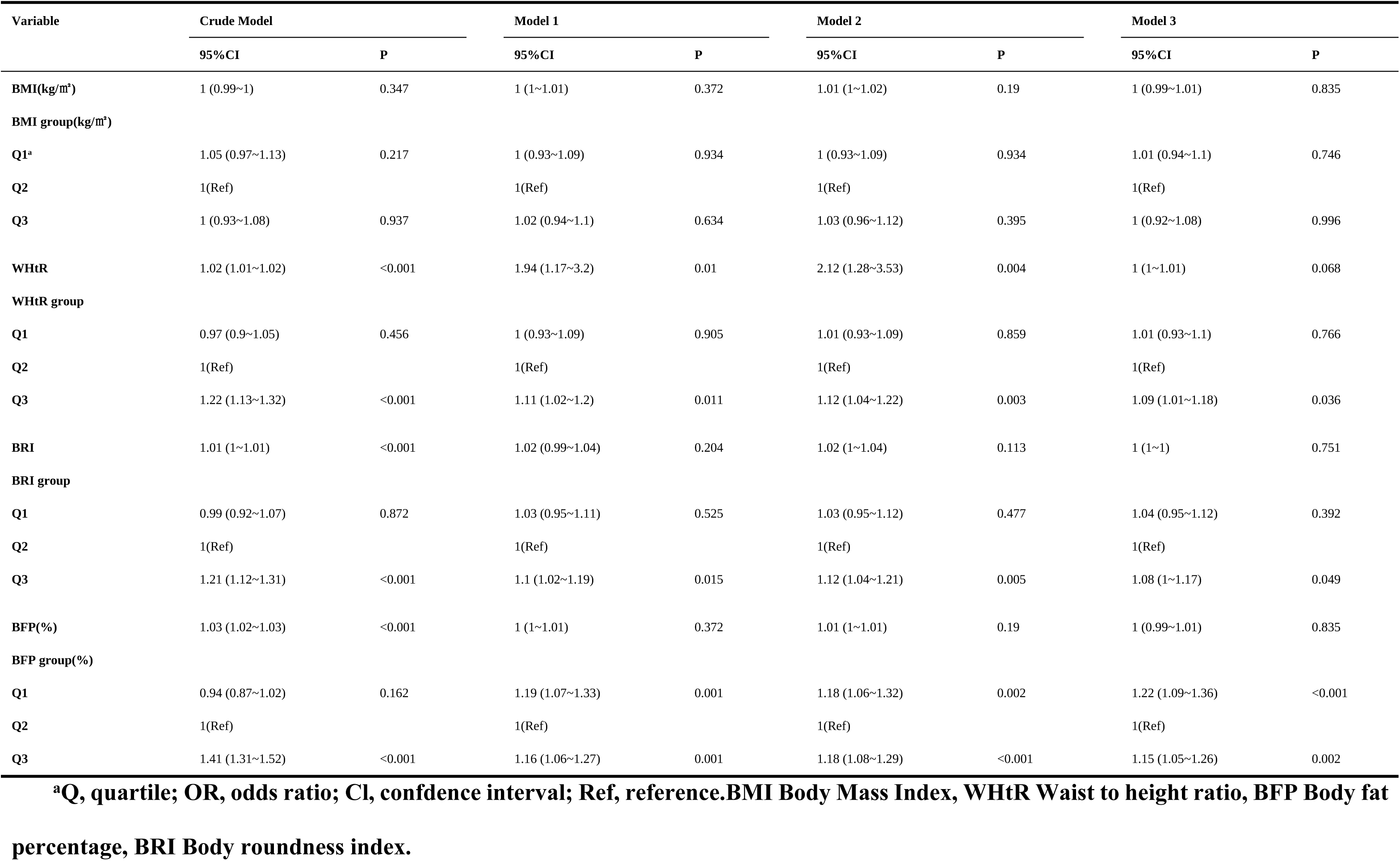
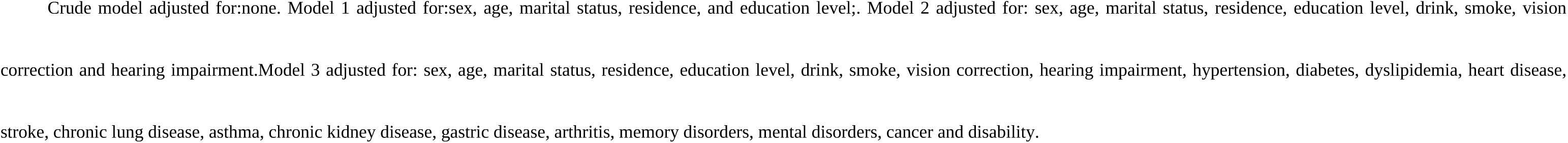
Association between obesity measurement indices and fall risk in different models.

| Variable | Crude Model |  | Model 1 |  | Model 2 |  | Model 3 |  |
| --- | --- | --- | --- | --- | --- | --- | --- | --- |
|  | 95%CI | P | 95%CI | P | 95%CI | P | 95%CI | P |
| **BMI(kg/m<sup>2</sup>)** | 1 (0.99~1) | 0.347 | 1 (1~1.01) | 0.372 | 1.01 (1~1.02) | 0.19 | 1 (0.99~1.01) | 0.835 |
| **BMI group(kg/m<sup>2</sup>)** |  |  |  |  |  |  |  |  |
| Q1<sup>a</sup> | 1.05 (0.97~1.13) | 0.217 | 1 (0.93~1.09) | 0.934 | 1 (0.93~1.09) | 0.934 | 1.01 (0.94~1.1) | 0.746 |
| Q2 | 1(Ref) |  | 1(Ref) |  | 1(Ref) |  | 1(Ref) |  |
| Q3 | 1 (0.93~1.08) | 0.937 | 1.02 (0.94~1.1) | 0.634 | 1.03 (0.96~1.12) | 0.395 | 1 (0.92~1.08) | 0.996 |
| **WHtR** | 1.02 (1.01~1.02) | <0.001 | 1.94 (1.17~3.2) | 0.01 | 2.12 (1.28~3.53) | 0.004 | 1 (1~1.01) | 0.068 |
| **WHtR group** |  |  |  |  |  |  |  |  |
| Q1 | 0.97 (0.9~1.05) | 0.456 | 1 (0.93~1.09) | 0.905 | 1.01 (0.93~1.09) | 0.859 | 1.01 (0.93~1.1) | 0.766 |
| Q2 | 1(Ref) |  | 1(Ref) |  | 1(Ref) |  | 1(Ref) |  |
| Q3 | 1.22 (1.13~1.32) | <0.001 | 1.11 (1.02~1.2) | 0.011 | 1.12 (1.04~1.22) | 0.003 | 1.09 (1.01~1.18) | 0.036 |
| **BRI** | 1.01 (1~1.01) | <0.001 | 1.02 (0.99~1.04) | 0.204 | 1.02 (1~1.04) | 0.113 | 1 (1~1) | 0.751 |
| **BRI group** |  |  |  |  |  |  |  |  |
| Q1 | 0.99 (0.92~1.07) | 0.872 | 1.03 (0.95~1.11) | 0.525 | 1.03 (0.95~1.12) | 0.477 | 1.04 (0.95~1.12) | 0.392 |
| Q2 | 1(Ref) |  | 1(Ref) |  | 1(Ref) |  | 1(Ref) |  |
| Q3 | 1.21 (1.12~1.31) | <0.001 | 1.1 (1.02~1.19) | 0.015 | 1.12 (1.04~1.21) | 0.005 | 1.08 (1~1.17) | 0.049 |
| **BFP(%)** | 1.03 (1.02~1.03) | <0.001 | 1 (1~1.01) | 0.372 | 1.01 (1~1.01) | 0.19 | 1 (0.99~1.01) | 0.835 |
| **BFP group(%)** |  |  |  |  |  |  |  |  |
| Q1 | 0.94 (0.87~1.02) | 0.162 | 1.19 (1.07~1.33) | 0.001 | 1.18 (1.06~1.32) | 0.002 | 1.22 (1.09~1.36) | <0.001 |
| Q2 | 1(Ref) |  | 1(Ref) |  | 1(Ref) |  | 1(Ref) |  |
| Q3 | 1.41 (1.31~1.52) | <0.001 | 1.16 (1.06~1.27) | 0.001 | 1.18 (1.08~1.29) | <0.001 | 1.15 (1.05~1.26) | 0.002 |

<sup>a</sup>Q, quartile; OR, odds ratio; CI, confidence interval; Ref, reference. BMI Body Mass Index, WHtR Waist to height ratio, BFP Body fat percentage, BRI Body roundness index.

### Nonlinear interconnected

We conducted curve fitting analysis to explore the relationship between WHtR, BRI, BMI, BFP, and fall events. The results showed that WHtR, BRI, and BFP were significantly associated with fall events and exhibited nonlinear relationships (Figure 2). For WHtR, all overall P values are less than 0.05 (three are less than 0.001, and one is 0.012), and the nonlinear test P values are all less than 0.05 (0.001, 0.011, 0.004, and 0.023, respectively). BRI and BFP showed stronger significant associations with falls (P < 0.001). However, the overall association between BMI and fall events was not significant (all P values > 0.05), nor was the nonlinear relationship significant (all but one P value > 0.09), suggesting that BMI may be less effective than the other three indicators in predicting fall events.

**Fig 2.**
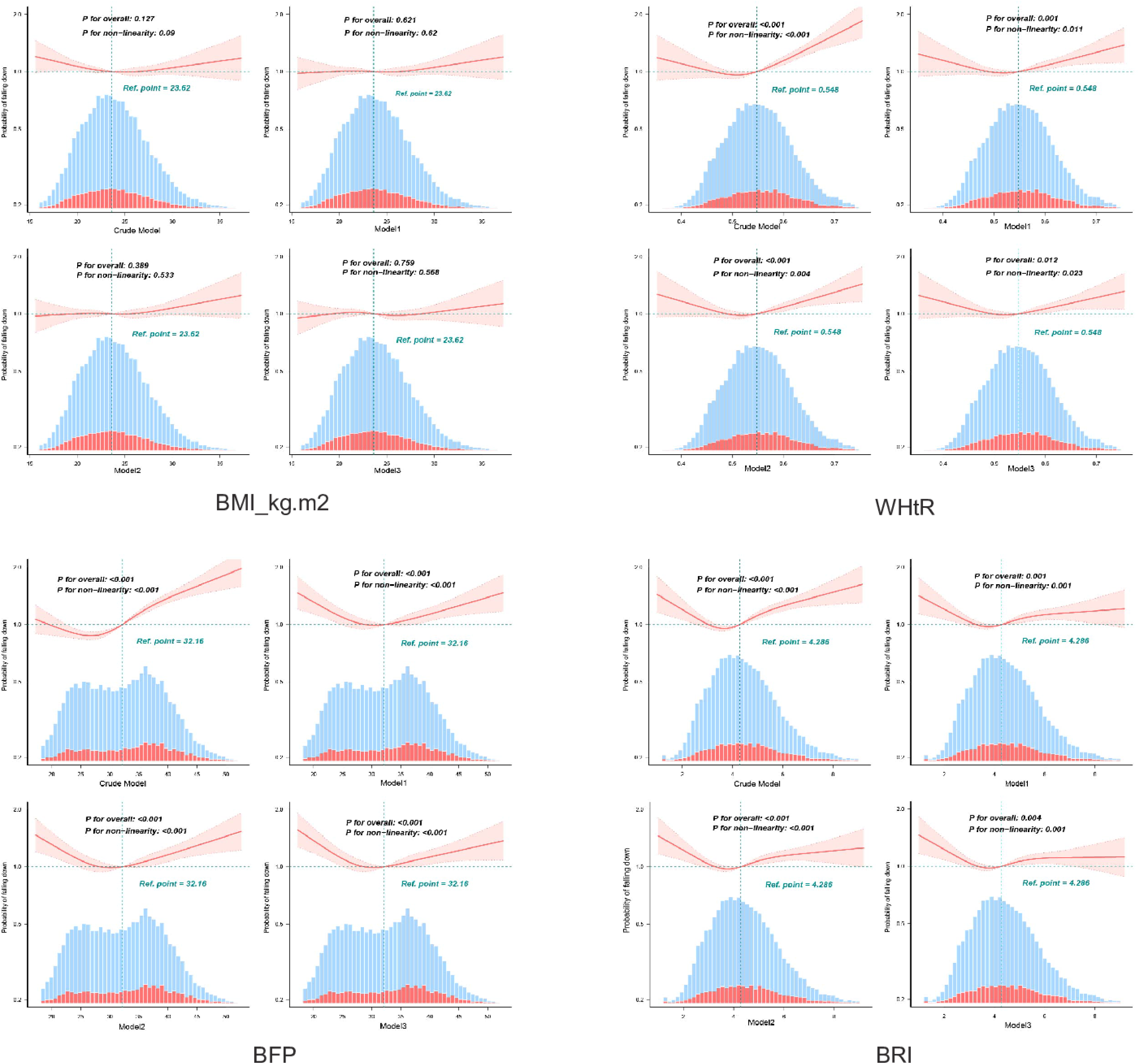
The RCS curve of the association between obesity measurement indices and fall

Using multivariable logistic regression models and smooth curve fitting, we observe nonlinear relationships between WHtR, BRI, BFP, and fall events. The data are fitted to a piecewise multivariable logistic regression model, which identifies two distinct slopes. We then apply a two-stage model to assess the associations between these three obesity measures and fall events (Table 3). WHtR exhibited an inflection point at 0.504 (95% CI: 0.502–0.506). Below this threshold, every 0.1 increment was associated with a 95% reduction in fall risk (OR = 0.05, 95% CI: 0.004–0.592; P = 0.018). Above the threshold, risk did not significantly increase (OR = 2.152, 95% CI: 0.976–4.748; P = 0.058). Although nonlinearity is statistically significant (P = 0.011), the post-threshold trend lacks statistical significance. The BRI inflection point is at 3.055 (95% CI: 2.92-3.19). Each 0.1 increase below this threshold reduces the risk of falls by 23.6% (OR = 0.764, 95% CI: 0.64-0.912; P = 0.003), while each 0.1 increase above this threshold increases the risk by 3.5% (OR = 1.035, 95% CI: 1.002-1.069; P = 0.039). Nonlinearity is statistically robust (likelihood ratio test: P < 0.001). The critical threshold for BFP is 28.575% (95% CI: 28.374-28.776). Below this inflection point, each 1% increase in BFP reduces the risk of falls by 3.2% (OR = 0.968, 95% CI: 0.946-0.99; P = 0.005). Conversely, above the threshold, each 1% increase in BFP increases the risk of falls by 1.3% (OR = 1.013, 95% CI: 1.002-1.024; P = 0.016). Likelihood ratio tests confirm that the nonlinear model fits better than the linear model (P < 0.001). Strong evidence of nonlinear dynamics was demonstrated for all three metrics by likelihood ratio tests (BFP: P < 0.001, BRI: P < 0.001, WHtR: P = 0.011) and quadratic nonlinearity tests (all P < 0.05). In contrast, BMI does not show significant nonlinear or linear associations with fall risk in preliminary analyses (all P > 0.05), justifying its exclusion from the piecewise modeling.

**Table 3.** Threshold effect analysis of obesity measurement indices on fall rick.

| Variable | Adjusted Model |  |  |  |  |  |
| --- | --- | --- | --- | --- | --- | --- |
|  | WHtR |  | BRI |  | BFP |  |
|  | OR (95%CI) | P value | OR (95%CI) | P value | OR (95%CI) | P value |
| **Inflection Point** | 0.504 (0.502 ,0.506) |  | 3.055 (2.92 ,3.19) |  | 28.575 (28.374 ,28.776) |  |
| **<Inflection Point** | 0.05 (0.004-0.592) | 0.0176 | 0.764 (0.64-0.912) | 0.0028 | 0.968 (0.946-0.99) | 0.0052 |
| **≥Inflection Point** | 2.152 (0.976-4.748) | 0.0576 | 1.035 (1.002-1.069) | 0.0387 | 1.013 (1.002-1.024) | 0.0158 |
| **Likelihood Ratio test** |  | 0.011 |  | <0.001 |  | <0.001 |

**Note: BMI Body Mass Index, WHtR Waist to height ratio, BFP Body fat percentage, BRI Body roundness index.**

### Subgroup analysis

Subgroup analysis (Figure 3) showed that sex, age, residence, and arthritis significantly modified the association between obesity and falls (interaction P<0.05). Specifically, a high WHtR was significantly associated with an increased risk of falls among women (OR=2.83, 95% CI: 1.45–5.53), individuals aged >60 years (OR=2.45, 95% CI: 1.23–4.88), rural residents (OR=2.77, 95% CI: 1.42–5.40), and patients with arthritis (OR=2.17, 95% CI: 1.01–4.63). No significant association was observed among men, individuals aged ≤59 years, urban residents, or those without arthritis. Although BFP and BRI showed statistical interaction in arthritis and other subgroups (P<0.05), their effect sizes were not clinically significant, as the 95% CIs of all ORs included 1 (see annex).

**Fig 3.**
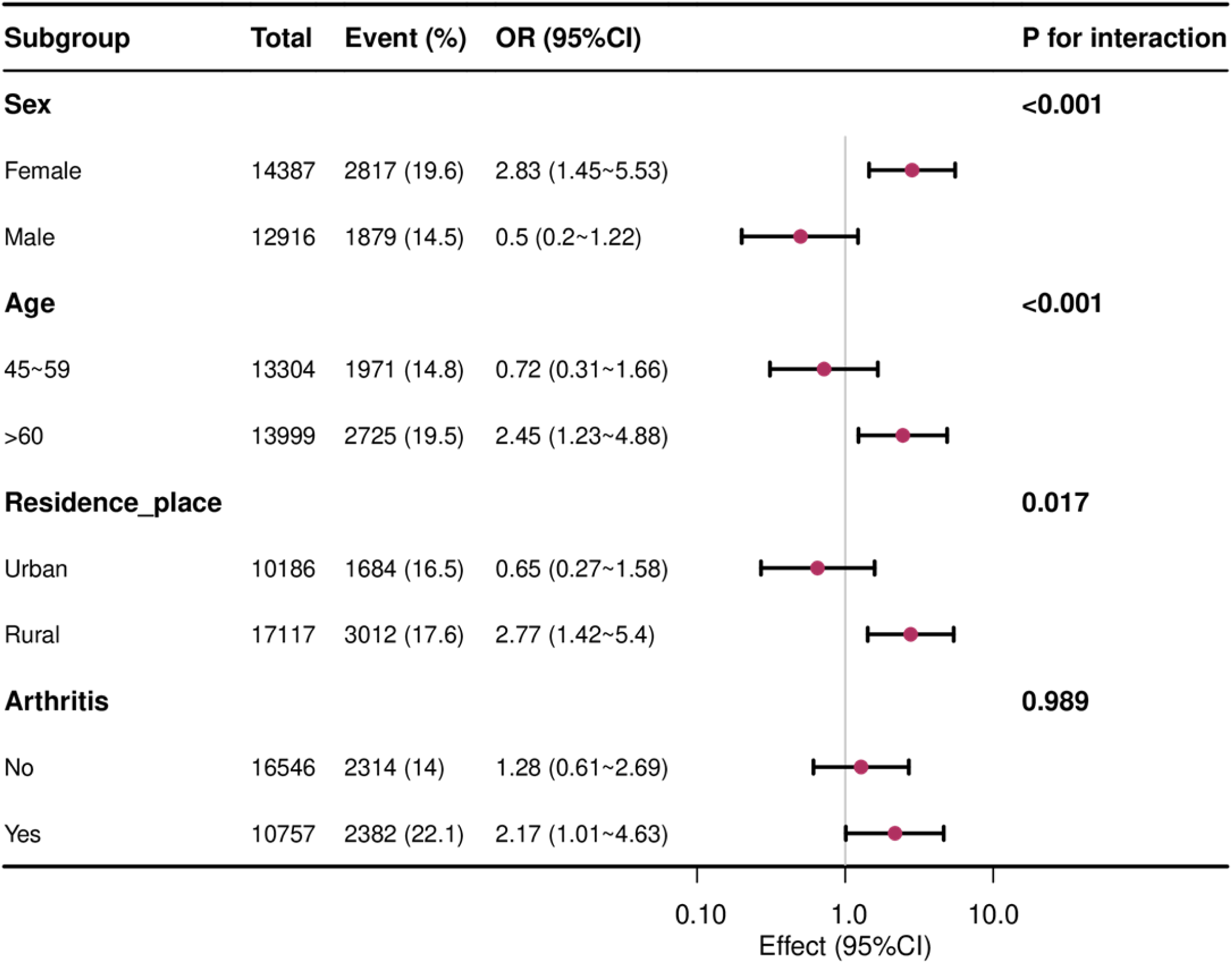
Associations between waist height ratio (WHtR) and fall rick in different subgroups

## Discussion

This cross-sectional study used data from the China Longitudinal Study of Health and Retirement (CHARLS) to systematically examine the nonlinear relationships and diverse mechanisms linking body mass index (BMI), body roundness index (BRI), body fat percentage (BFP), and waist height ratio (WHtR) with fall risk in older adults. The main findings showed that WHtR, BRI, and BFP—key indicators of central obesity and body composition—had significant nonlinear dose-response relationships with fall risk compared to BMI, with clear inflection points at 0.5 for WHtR, 3.06 for BRI, and 28.6% for BFP. The stepwise regression model further confirmed that fall risk decreased by 3.2% (OR = 0.968, P = 0.005) for every unit increase in BFP below the inflection point in the fully adjusted model, but increased after exceeding the inflection point (OR = 1.013, P = 0.016). Similarly, BRI exhibited a significant U-shaped nonlinear relationship with fall events, highlighting the limitation of the traditional BMI’s linear assumption. Compared with the overall obesity level reflected by BMI, WHtR, BRI, and BFP are more directly related to fat distribution in obese individuals. Among these indicators, visceral fat accumulation may directly or indirectly lead to muscle function decline [20]. Moreover, existing evidence shows that body composition imbalance is a key pathological basis: increased fat-muscle ratio (FMR) has been confirmed as the strongest predictor of muscle function decline and is significantly associated with fall risk [21]. Another study also revealed a significant negative correlation between fat-muscle ratio and walking speed and grip strength, suggesting that increasing fat ratio weakens muscle strength and coordination, thereby increasing the risk of loss of homeostasis [22]. Specifically, the observed risk increase beyond the inflection point aligns with previous studies. Laboratory experiments found that obese individuals have significantly higher slip and fall rates than non-obese individuals, due to impaired dynamic balance and failed recovery strategies [23]. Moreover, a large meta-analysis including 31 studies with 1,758,694 participants confirmed this association, showing a 16% increased fall risk among obese adults over 60 years old (RR = 1.16, 95% CI: 1.07–1.26) [24]. Obese individuals, especially those with abdominal obesity combined with low grip strength, had a 4.1-fold increased risk of fall injury compared to normal individuals (OR = 4.10, 95% CI 2.81–5.98), highlighting the synergistic harm of muscle-fat metabolism interaction [25]. Before the inflection point, the reduction in risk may be explained by the positive effect of appropriate fat on fall prevention. Undernourishment also poses a significant fall risk factor in the elderly. For example, a prospective cohort study of community-dwelling elderly found that high malnutrition risk was associated with a 66% increase in the probability of falls within the next year [26].In addition, malnourished patients were significantly more likely to experience falls than non-malnourished patients in hospital settings, particularly among patients over 80 years of age [27]. The risk of malnutrition is associated with several age-related syndromes, such as polytherapy, falls, frailty, insomnia, and depression [28]. Moreover, low lean body mass and high body fat percentage have also been shown to be directly associated with falls. This suggests that changes in body composition should be considered when assessing the functional capacity and safety of older adults [29].

The association between WHtR and falls varied significantly by gender and residence. Specifically, rural women with high WHtR had a significantly higher risk of falls, whereas urban men showed no significant difference. This difference is due to a biosocial interaction mechanism: the decline in estrogen in postmenopausal women accelerates muscle loss and central adiposity [11,30]. In rural areas, the negative effects of obesity are amplified by insufficient medical resources and high-risk activity environments, such as uneven roads [31], which is consistent with Mitchell et al.’s study on the high risk of obesity in women [32]. A cohort study in Brazil further confirmed that female obesity, dominated by hip fat, with a body fat percentage (BFP) >42% doubled the risk of falls compared with non-obese individuals (RR=2.09, 95% CI 1.13–3.87). In contrast, male obesity, dominated by abdominal fat, with the same BMI did not have a statistically different risk (RR=1.26, 95% CI 0.64–2.50) [33]. Available evidence also suggests that BFP ≥28% independently predicts fracture risk (OR=1.71, 95% CI 1.08–2.71), providing quantitative evidence for secondary prevention [34]. Based on this, a dual-track strategy is recommended for clinical practice. Specifically, BRI≥3.0 and BFP≥28% should be incorporated into the elderly health screening system as fall risk assessment thresholds. Differentiated interventions are also necessary, such as resistance training for women to reverse muscle function inhibition, and a combination of environmental modifications and obesity management education in rural areas.Future prospective studies are needed to verify the applicability of these threshold values across populations.

CHARLS data are collected and screened according to standardized procedures to ensure accuracy and consistency of results. The results are reliable because they are based on a large community sample. Regression analysis accounts for important confounding factors, providing a comprehensive understanding of associations. However, it is not directly comparable to mechanistic studies, as they address different aspects of research.

However, the following limitations need to be carefully considered. First, although cross-sectional designs control for confounding through multiple imputation and multivariate adjustments, including demographics, lifestyle habits, and chronic diseases, it is still difficult to establish causality. Second, there is recall bias in the self-reporting of falls and a lack of objective tests of balance function, such as center of pressure displacement measurements, to directly verify biomechanical mechanisms. Third, although the sample is representative of the whole country, it does not cover ethnic minority regions and vulnerable elderly groups, such as those over 80 years old, which limits the generalizability of the findings. Since the study participants were exclusively from China, caution is required when generalizing the results to other countries or ethnic groups worldwide. To further validate our results, large-scale cohort and mechanistic studies are needed for validation.

## Conclusion

This study demonstrates that WHtR, BRI, and BFP are superior to BMI in predicting fall risk among older Chinese adults. The observed nonlinear relationships and identified thresholds (BRI ≥ 3.0, BFP ≥ 28%) may guide future fall prevention strategies. Incorporating these indices into clinical assessments could enhance early risk identification and targeted interventions.

## Data Availability

All data files are available from the CHARLS database(https://charls.pku.edu.cn/)

## Acknowledgments

We appreciate the China Health and Retirement Longitudinal Study team for providing data and training in using the datasets.

## Author contributions

Zihao Liu. designed the study, analyzed the data, and wrote the manuscript. Jiahe Zhang. summarize and analyze data analyzed the data. YongJun Li. revised the manuscript. All authors read and approved the final manuscript.

## Data availability statement

All raw data and code are available upon request.

## Ethics statement

The real data is publicly available, thus ethics approval was not required.

## References

1. Hatamabadi HR, Sum S, Tabatabaey A, Sabbaghi M. Emergency department management of falls in the elderly: A clinical audit and suggestions for improvement. Int Emerg Nurs. 2016;24:2–8.

2. Kalula SZ, Ferreira M, Swingler GH, Badri M. Risk factors for falls in older adults in a South African Urban Community. BMC Geriatr. 2016;16:51.

3. Liu SW, Obermeyer Z, Chang Y, Shankar KN. Frequency of ED revisits and death among older adults after a fall. Am J Emerg Med. 2015;33:1012–8.

4. Oseni TIA, Ibharokhonre AO, Olawumi AL, Iyalomhe ES, Adebayo CU, Adewuyi BO, Fuh FN. Association between obesity, physical activity and falls among elderly patients attending the family medicine clinics of a teaching hospital in Southern Nigeria. BMC Geriatr. 2025;25:93.

5. Liu M, Kang N, Wang D, Mei D, Wen E, Qian J, Chen G. Analysis of lower extremity motor capacity and foot plantar pressure in overweight and obese elderly women. Int J Environ Res Public Health. 2023;20:3112.

6. Jeon BJ. The effects of obesity on fall efficacy in elderly people. J Phys Ther Sci. 2013;25:1485–9.

7. Siqueira FV, Facchini LA, Silveira DS, Piccini RX, Tomasi E, Thumé E, Silva SM, Dilélio A. Prevalence of falls in elderly in Brazil: a countrywide analysis. Cad Saude Publica. 2011;27:1819–26.

8. Lee JJ, Hong DW, Lee SA, Soh Y, Yang M, Choi KM, Won CW, Chon J. Relationship between obesity and balance in the community-dwelling elderly population: a cross-sectional analysis. Am J Phys Med Rehabil. 2020;99:65–70.

9. Rosic G, Milston AM, Richards J, Dey P. Fear of falling in obese women under 50 years of age: a cross-sectional study with exploration of the relationship with physical activity. BMC Obes. 2019;6:7.

10. Frankenfield DC, Rowe WA, Cooney RN, Smith JS, Becker D. Limits of body mass index to detect obesity and predict body composition. Nutrition. 2001;17:26–30.

11. Dam TV, Dalgaard LB, Ringgaard S, Johansen FT, Bisgaard Bengtsen M, Mose M, Lauritsen KM, Ørtenblad N, Gravholt CH, Hansen M. Transdermal estrogen therapy improves gains in skeletal muscle mass after 12 weeks of resistance training in early postmenopausal women. Front Physiol. 2021;11:596130.

12. Gao X, Wang L, Shen F, Ma Y, Fan Y, Niu H. Dynamic walking stability of elderly people with various BMIs. Gait Posture. 2019;68:168–73.

13. Handrigan GA, Maltais N, Gagné M, Lamontagne P, Hamel D, Teasdale N, Hue O, Corbeil P, Brown JP, Jean S. Sex-specific association between obesity and self-reported falls and injuries among community-dwelling Canadians aged 65 years and older. Osteoporos Int. 2017;28:483–94.

14. Huang L, Shen X, Zou Y, Wang Y. Effects of BMI and grip strength on older adults’ falls—a longitudinal study based on CHARLS. Front Public Health. 2024;12:1415360.

15. Zhao Y, Hu Y, Smith JP, Strauss J, Yang G. Cohort profile: the China Health and Retirement Longitudinal Study (CHARLS). Int J Epidemiol. 2014;43:61–8.

16. Du G, Fan Z, Fan K, Liu H, Zhang J, Li D, Yan L, Jiu J, Li R, Li X, Li S, Jia L, Liu H, Ren Y, Liu X, Li JJ, Wang B. Risk-stratified lifetime risk and incidence of hip fracture and falls in middle-aged and elderly Chinese population: The China health and retirement longitudinal study. J Orthop Translat. 2025;50:174–84.

17. Guo T, Zhang F, Xiong L, Huang Z, Zhang X, Wan J, Mo J. Association of handgrip strength with hip fracture and falls in community-dwelling middle-aged and older adults: A 4-year longitudinal study. Orthop Surg. 2024;16:1051–63.

18. Thomas DM, Bredlau C, Bosy-Westphal A, Mueller M, Shen W, Gallagher D, Maeda Y, McDougall A, Peterson CM, Ravussin E, Heymsfield SB. Relationships between body roundness with body fat and visceral adipose tissue emerging from a new geometrical model. Obesity (Silver Spring). 2013;21(2):2264–71.

19. Lv H, Wang Y, Zhang G, Wang X, Hu Z, Chu Q, Zhou Y, Yang Y, Jiang T, Wang J. Association between obesity measurement indexes and symptomatic knee osteoarthritis among the Chinese population: analysis from a nationwide longitudinal study. BMC Musculoskelet Disord. 2024;25:986.

20. Rico-Martín S, Calderón-García JF, Sánchez-Rey P, Franco-Antonio C, Martínez Alvarez M, Sánchez Muñoz-Torrero JF. Effectiveness of body roundness index in predicting metabolic syndrome: a systematic review and meta-analysis. Obes Rev. 2020;21(5):e13023.

21. Kao TW, Peng TC, Chen WL, Han DS, Chen CL, Yang WS. Impact of adiposity on muscle function and clinical events among elders with dynapenia, presarcopenia and sarcopenia: a community-based cross-sectional study. Aging (Albany NY). 2021;13:7247–58.

22. Chao YP, Chen WL, Peng TC, Wu LW, Liaw FY, Kao TW. Examining the association between muscle mass, muscle function, and fat indexes in an elderly population. Nutrition. 2022;83:111071.

23. Allin LJ, Wu X, Nussbaum MA, Madigan ML. Falls resulting from a laboratory-induced slip occur at a higher rate among individuals who are obese. J Biomech. 2016;49:678–83.

24. G R Neri S, S Oliveira J, B Dario A, M Lima R, Tiedemann A. Does obesity increase the risk and severity of falls in people aged 60 years and older? A systematic review and meta-analysis of observational studies. J Gerontol A Biol Sci Med Sci. 2020;75:952–60.

25. Dowling L, McCloskey E, Cuthbertson DJ, Walsh JS. Dynapenic abdominal obesity as a risk factor for falls. J Frailty Aging. 2023;12:37–42.

26. Eckert C, Gell NM, Wingood M, Schollmeyer J, Tarleton EK. Malnutrition risk, rurality, and falls among community-dwelling older adults. J Nutr Health Aging. 2021;25:624–7.

27. Eglseer D, Hoedl M, Schoberer D. Malnutrition risk and hospital-acquired falls in older adults: A cross-sectional, multicenter study. Geriatr Gerontol Int. 2020;20:348–53.

28. Sucuoglu Isleyen Z, Besiroglu M, Yasin AI, Simsek M, Topcu A, Smith L, Akagunduz B, Turk HM, Soysal P. The risk of malnutrition and its clinical implications in older patients with cancer. Aging Clin Exp Res. 2023;35:2675–83.

29. Hashim NNA, Mat S, Myint PK, Kioh SH, Delibegovic M, Chin AV, Kamaruzzaman SB, Hairi NN, Khoo SPK, Tan MP. Association between weight and body composition changes with falls risk in the Malaysian Elders Longitudinal Research (MELoR) study. BMJ Open. 2024;14:e087358.

30. Mandelli A, Tacconi E, Levinger I, Duque G, Hayes A. The role of estrogens in osteosarcopenia: from biology to potential dual therapeutic effects. Climacteric. 2022;25:81–7.

31. Norman K, Burrows L, Chepulis L, Mullins H, Lawrenson R. ’They’re all individuals, none of them are on the same boat’: barriers to weight management in general practice from the rural nurse perspective. Prim Health Care Res Dev. 2023 Jul 31;24:e50.

32. Mitchell RJ, Lord SR, Harvey LA, Close JC. Associations between obesity and overweight and fall risk, health status and quality of life in older people. Aust N Z J Public Health. 2014;38:13–8.

33. Neri GR, Tiedemann A, Gadelha AB, Lima RM. Body fat distribution in obesity and the association with falls: a cohort study of Brazilian women aged 60 years and over. Maturitas. 2020;139:64–8.

34. Gandham A, Zengin A, Bonham MP, Winzenberg T, Balogun S, Wu F, Aitken D, Cicuttini F, Ebeling PR, Jones G, Scott D. Incidence and predictors of fractures in older adults with and without obesity defined by body mass index versus body fat percentage. Bone. 2020;140:115546.

